# Pandemic risk in the Shared Socioeconomic Pathways

**DOI:** 10.64898/2026.07.29.26359250

**Authors:** Torre E. Lavelle, Cecilia A. Sánchez, Marina Andrijevic, Daniel J. Becker, Rory Gibb, Gregg S. Gonsalves, Zoe O’Donoghue, Shonali Pachauri, Laura M. Pereira, Timothée Poisot, Sadie J. Ryan, Stephanie N. Seifert, Charles Whittaker, Colin J. Carlson

## Abstract

For over a decade, the Shared Socioeconomic Pathways (SSPs) have served as the principal framework for quantitative modeling of the socioeconomic dimensions of global environmental change. The SSP scenarios describe many of the ecological and social processes thought to shape pandemic risk, including the emergence of novel pathogens (accelerated by processes such as deforestation, livestock intensification, and land-use change) and their subsequent spread (mediated by factors such as inequality, human mobility, and health system capacity). However, the SSP framework has not been widely incorporated into pandemic risk assessment. Here, we assess how pandemic risk is embedded in the SSP framework, and find that the framework captures most of the social-environmental drivers of pathogen spillover, and many of the social-economic drivers of pandemic spread and impacts. Because climate change and pandemics share many drivers and risk factors—including ecosystem degradation, animal agriculture, and weak governance—SSP scenarios characterized by higher barriers to climate adaptation also generally imply lower chances of outbreak containment, and greater pandemic impacts on vulnerable populations. Pandemic risk is therefore lowest in SSP1 and highest in SSP3, but SSP5 shows that frequent spillover and effective containment can coexist. These findings suggest that pandemic risk can be understood as part of a broader polycrisis, linking climate change, biodiversity loss, and global health. We suggest that new scenario extensions, or entirely novel frameworks, will ultimately be needed to capture possible shifts in the global health landscape; however, in the meantime, scenario frameworks from the environmental sciences could be valuable tools for initiatives to quantify future pandemic risks.

## Introduction

While infectious disease outbreaks are naturally occurring phenomena, the recent rise of emerging infectious diseases is generally understood to be the result of social, economic, and environmental change^1–5^. The social-ecological drivers of disease emergence include factors that mediate disease dynamics in nature (e.g., deforestation and climate change) and interfaces that allow zoonotic spillover (animal-to-human pathogen transmission) to occur (e.g., livestock production and wildlife trade). Once a novel pathogen begins to spread in human populations, the trajectory of a pandemic (an epidemic with global spread and impact; see **Box 1**) is shaped by population characteristics (e.g., age structure), poverty and economic inequality (within and across countries), global travel and trade (e.g., multi-scale mobility), health system capacity and preparedness, individual and institutional behavior (e.g., adherence to public health measures and the provision of social safety nets), and political, governance, and legal factors (e.g., vaccine nationalism or use of restrictive measures like lockdowns). All of these factors are constantly shifting, and could follow a wide range of trajectories over the coming century.

#### Box 1. Glossary. These definitions are not exclusive to other interpretations of these terms in the literature, but provide a standardized jumping off point for this study.

- **Emergence:** An expansion of pathogen host range and geographic range with relevance to human health. This process can be gradual or sudden, and can occur naturally, but is often accelerated by ecological and social drivers. In the case of pandemic risk, disease emergence often follows a stepwise process of zoonotic spillover (animal-to-human transmission) followed by epidemic or potentially pandemic spread (human-to-human transmission).
- **Integrated assessment model (IAM):** Coupled human–natural system models used in climate change research that simulate interactions among population, economic development, energy systems, agriculture, land use, and greenhouse gas emissions.
- **Pandemic:** An epidemic of an infectious disease with global spread and impact. The term itself is subjective, contested, and political; our definition is selected to make the most meaningful distinction between “epidemic” (which can be any acute, severe outbreak) and “pandemic.” Historically, pandemics have included bacterial diseases like cholera and plague, but most modern pandemics (and all pandemics that have started in the last fifty years) have been caused by viruses^5^. Usually, these viruses enter the human population from an animal reservoir, although other pathways (e.g., biosafety lapses) do exist.
- **Pandemic risk:** The long-run frequency and impact of pandemics on human societies, economies, and ecosystems. That risk is the product of several component processes, including the risk of zoonotic spillover; the risk of pandemic spread; risk multipliers that increase pandemic impacts or distribute them unequally; and the effect of interventions targeting each stage. Each of these risks may be further disaggregated (e.g., following the hazard-exposure-vulnerability framework^6^).
- **Representative Concentration Pathways (RCPs):** A scenario framework that has been widely used in climate change research. The RCPs consist of four (and occasionally more) scenarios describing plausible trajectories of greenhouse gas concentrations through 2100 (and sometimes beyond). Currently, the research community is in the process of departing from the RCPs, in favor of new emissions-, rather than concentrations-, based pathways^7^.
- **Scenario extensions:** Narratives or models that are produced to capture aspects or scales of social and environmental change not captured in a scenario framework. Here, these are usually quantitative or qualitative extensions of the original SSP narratives.
- **Scenario framework:** A standardized set of scenarios used in scientific research; here, generally referring to a set of descriptive and quantitative assumptions about global environmental and social change over at least the next century.
- **Scenario narratives**: A set of descriptions or storylines that underpin a scenario framework.
- **Shared Socioeconomic Pathways (SSPs):** A scenario framework that has been widely used in the last decade of research on climate change and sustainable development. The SSPs consist of five scenarios that describe a range of worlds with low, medium, or high challenges to climate change mitigation and adaptation.
- **Zoonotic spillover:** Animal-to-human pathogen transmission (abbreviated as simply spillover throughout).

In the environmental sciences (especially among researchers studying the twin challenges of climate change and biodiversity loss), this uncertainty about future social and environmental change is often addressed using a small number of standardized scenarios. These scenario frameworks create a common touchpoint for possible, or sometimes preferable, visions of planetary futures. When articulated in quantitative terms, they allow researchers to project impacts of global change on human societies and the environment, and even clearly define the benefits or harms of specific policy decisions^8,9^. By standardizing these scenarios, researchers can more readily compare modeling outputs and assess uncertainty.

Over the last few decades, there have been several generations of scenarios used in climate change research; since 2011, the Coupled Model Intercomparison Project (CMIP)—the global organizing network for climate modeling—has used the Representative Concentration Pathways (RCPs)^10,11^ (but see **Box 1**). The Shared Socioeconomic Pathways (SSPs) were introduced in 2014 to complement the narrowly greenhouse gas-focused RCPs, with the intention to represent the socioeconomic dynamics that could underlie different emissions pathways, and for use in climate change impact studies that need estimates of future population size and structure, economies, urbanization, and similar variables. The SSPs consist of five scenarios that describe a range of worlds with low, medium, or high challenges to climate change mitigation and adaptation^12,13^. When used together, the RCPs and SSPs create a matrix of scenarios with a range of emissions trajectories and underlying socio-economic development, including the representation of barriers to climate change adaptation and mitigation, with some combinations ruled out due to internal inconsistency (e.g., sustainable socioeconomic development is incompatible with the highest emissions forcing).

In public health, the SSP–RCP scenario framework has been used to project a wide range of future health harms of climate change^14–16^, including impacts on infectious disease dynamics. So far, this work has focused more on endemic diseases like malaria and dengue fever, and less on potential pandemic threats^17^. However, climate change is likely to shift the geographic distributions of many wildlife hosts of pathogens with pandemic potential, and changes in weather may also mediate pathogen transmission in some systems^18–22^. Climate change will also affect pandemic vulnerability indirectly through its effects on poverty, conflict, infrastructure, and health systems^23–25^. Evidence of these linkages is still mostly speculative, and there is currently no consensus that climate change constitutes a dominant or generalizable driver of pandemic risk^3,5^. However, there is general agreement that the connections between climate change and pandemic risk should be explored in more depth.

Up to this point, pandemic risk assessment has primarily engaged with the SSP–RCP framework as a tool for studying climate change impacts^19,26^. However, the SSPs are themselves a rich text, describing long-term visions of social, economic, environmental, and institutional conditions that would all shape pandemic risk across the pandemic emergence process. Each SSP storyline was originally developed as a narrative, and quantified with basic models that describe population dynamics (including age structure and education level), economic growth, and urbanization^27–29^. These projections, and the underlying narratives, are used as inputs by integrated assessment models (IAMs), a class of models that explore trajectories for future emissions and options to reduce them. Some studies have also developed qualitative and quantitative extensions of the SSP framework, which address aspects or scales of social and environmental change not captured by the original narratives, including health system transformation and non-communicable disease burden^30–33^—but so far, not pandemic risk. A handful of studies have used the SSP or SSP–RCP scenarios to project how future land use change could redistribute animal reservoirs or vectors of zoonotic viruses and alter opportunities for zoonotic spillover and onward spread^18,19,26,34^. However, other facets of social and ecological transformation captured in the SSPs—such as agricultural intensification, improvements in sanitation, and geopolitical fragmentation—also have immediate, and so far unexplored, relevance to pandemic risk

Here, we ask how pandemic risk is envisioned in, embedded in, or absent from the SSPs. Our analysis is organized around a stepwise framework of pandemic emergence that is widely used in disease ecology and pandemic risk assessment^35–37^. Throughout this paper, we use “pandemic risk” as an umbrella concept encompassing the entire pandemic emergence process—from pathogen spillover and the establishment of sustained human-to-human transmission to epidemic and pandemic spread, societal response, and the resulting health, social, and economic impacts. In this framework, pandemic risk is shaped by a sequence of linked processes: first, ecological and social conditions create opportunities for pathogen exposure and spillover from animals to humans; second, pathogen, host, and population-level factors determine whether spillover sparks sustained human-to-human transmission; third, mobility, demography, inequality, and health system capacity shape whether local outbreaks expand into epidemics or pandemics; and finally, surveillance, governance, medical countermeasures, and social protections determine the scale and distribution of pandemic impacts. This framework allows us to distinguish drivers of spillover from drivers of onward spread, response, and impact, while recognizing that all contribute to overall pandemic risk.

First, we identify a wide-ranging set of pandemic risk drivers across these stages and examine whether, and in how much detail, they are represented in the SSP narratives. We follow a similar qualitative approach to previous work exploring how the SSPs align with other aspects of social change, such as democratic governance and digital transformation^38–40^. Where possible, we also use quantitative data to explore the range of envisioned trajectories for key variables, such as livestock production and land use change. Based on this analysis, we ask whether the alternative socioeconomic and political futures represented by the SSPs, and their associated challenges to climate change mitigation and adaptation, also imply different trajectories of pandemic risk. Finally, we discuss the limitations of applying the SSP framework to pandemic risk assessment, and possible next steps for scenario uptake and development in this field.

## Methods

### The Shared Socioeconomic Pathways framework

The SSPs were originally developed to provide a common framework for exploring how alternative patterns of socioeconomic development set the difficulty of climate change mitigation and adaptation. The framework consists of five short narrative paragraphs describing contrasting futures for global demographic, economic, technological, institutional, and environmental change, ranging from sustainability-oriented development (SSP1) to middle of the road (SSP2), regional rivalry (SSP3), inequality (SSP4), and fossil-fueled development (SSP5). Importantly, the SSP narratives do not describe specific predictions about the future, or make assumptions about future climate change or its impacts. Instead, these five scenarios provide representative examples of alternative socioeconomic futures that can be coupled with greenhouse gas emissions trajectories to create cohesive visions of possible climate futures.

The five SSPs were intentionally selected to span a two-dimensional scenario space defined by the challenges societies face for climate change mitigation and adaptation^13^. SSP1–SSP3 represent worlds with low, medium, high challenges (respectively) to both mitigation and adaptation; while SSP4 and SSP5 illustrate contrasting combinations in which mitigation and adaptation challenges diverge. SSP4 combines relatively low barriers to mitigation with high barriers to adaptation because wealth, technological capacity, and institutional strength remain concentrated in high-income countries, while poorer regions continue to experience persistent poverty and environmental degradation. SSP5 combines low adaptation challenges with high mitigation challenges through rapid economic growth, technological progress, and continued reliance on fossil fuels. These two final scenarios are intentionally stylized and complete the two-dimensional SSP scenario space by representing combinations of mitigation and adaptation challenges that differ from the more monotonic progression represented by SSP1–SSP3.

The original SSP narratives have been translated into quantitative projections of a small set of core demographic and economic variables^27–29^, and then—with the use of IAMs—of other variables such as those related to land and energy use. Multiple IAMs have implemented the SSP framework, each using different model structures, assumptions, and representations of socioeconomic processes while remaining consistent with the overarching SSP narratives. The resulting projections are hosted by the International Institute for Applied Systems Analysis (IIASA) in the SSP Scenario Explorer database, which provides comparable estimates of demographic, economic, agricultural, and land-use variables across scenarios. A handful of studies have also developed quantitative extensions of the SSPs for other variables, such as civil conflict or gender inequality; these are also harmonized in the IIASA SSP Extensions Explorer. All external data we use in this study were accessed from these two databases.

### Identification of pandemic risk drivers

To assess how pandemic risk is represented in the SSP narratives and differs between scenarios, we first adopted a conceptual framework describing the pandemic emergence process, in which pandemic risk arises through sequential stages from pathogen spillover to pandemic spread, societal impacts, and public health response. Guided by this framework, we then developed a core set of social, ecological, institutional, and technological drivers known to be associated with pandemic risk (**Figure 1**). This list was not intended to be comprehensive, but captures the most important thematic elements in current literature on pandemic risk, particularly as they relate to environmental and demographic change.

**Figure 1.**
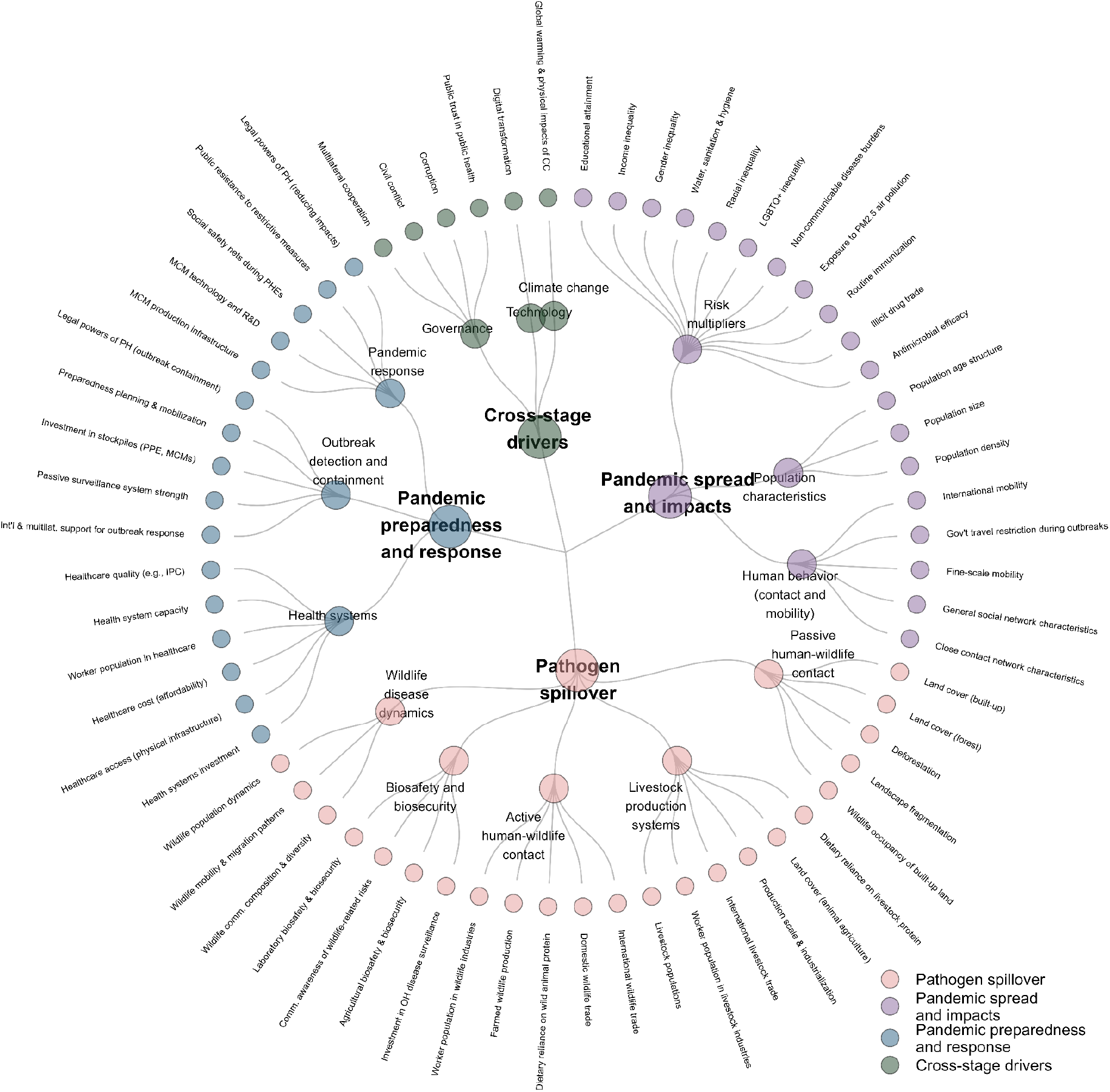
Pandemic risk drivers. Classification of 64 pandemic risk drivers identified through a purposive literature review and synthesis of existing frameworks in disease emergence, pandemic prevention, and preparedness (see Methods). Drivers are organized by their primary stage of action in the pandemic emergence process (spillover, pandemic spread and impacts, pandemic preparedness and response, or cross-cutting drivers) and grouped into thematic categories (e.g., wildlife disease dynamics, active human-wildlife contact, health systems, and governance). CC = climate change; IPC = infection prevention and control; LGBTQ = lesbian, gay, bisexual, transgender, and queer; MCMs = medical countermeasures; OH = One Health; PH = public health; PHEs = public health emergencies; PPE = personal protective equipment; R&D = research and development.

To identify these drivers, we first conducted a purposive literature review using Google Scholar to identify review articles and foundational publications on zoonotic spillover, pandemic spread and impacts, and pandemic prevention, preparedness, and response measures. We selected Google Scholar because its broad coverage captures interdisciplinary and grey literature that may not be indexed in discipline-specific databases, making it well suited to identifying conceptual frameworks and review articles across the environmental and health sciences^41,42^. Searches combined terms related to pandemic risk, including “zoonotic spillover,” “emerging infectious diseases,” “pandemic preparedness,” “pandemic prevention,” “disease emergence,” “land-use change,” “health systems,” and “surveillance.” We paid particular attention to review articles that have provided a comprehensive overview of scientific consensus on the drivers of disease emergence^3,5,43^. Candidate drivers were iteratively refined through discussion among the authors. We screened these studies to identify drivers of pandemic risk, which we defined as social, ecological, institutional, or technological processes that influence: i) the likelihood of pathogen spillover from animals to humans; ii) onwards human-to-human transmission; iii) the clinical, social, and economic burden of these infections; or iv) societal capacity to detect and respond to outbreaks.

This process generated an initial list of candidate drivers that we grouped into four stages of pandemic risk: pathogen spillover, pandemic spread and impacts, pandemic preparedness and response, and cross-stage drivers. Pathogen spillover drivers included processes that increase contact between humans, livestock, and wildlife or alter pathogen ecology (e.g., landscape fragmentation, international livestock trade). (Here, we focus on zoonotic spillover as the origin of human infection, because this represents the majority of both modern pandemics and emerging infectious disease outbreaks^1,5^; however, we include laboratory biosafety and biosecurity as another risk driver at the spillover stage, given that accidental infections particularly pose a non-zero risk^44^.) Drivers of pandemic spread and impacts included factors influencing human-to-human transmission or outcomes of infection (e.g., population density, income inequality). Pandemic preparedness and response drivers included institutional and health system characteristics shaping outbreak detection, surveillance, and containment (e.g., health systems investment, preparedness planning and mobilization). Finally, a handful of drivers are relevant across all three stages (e.g., climate change and multilateral cooperation).

This preliminary list of drivers was refined through iterative discussion among the authors, and a handful of drivers were revised, aggregated, split, or added based on expert opinion, in order to ensure each driver was well-supported, non-redundant, and sufficiently granular to map onto the SSP narratives. Through this combined literature review and expert synthesis, we identified a final list of 64 pandemic risk drivers.

### Mapping pandemic risk drivers onto the SSP framework

To map the drivers onto the SSP framework, we examined: the SSP narratives; the accompanying quantifications of core exogenous drivers (i.e., population change, including age and education; economic growth; and urbanization); IAMs quantifications; SSP narrative extensions, particularly related to health^30,31,33^; and studies that use non-IAM models to project change in other variables under the SSP scenarios (some of which are described as scenario extensions). Ambiguous cases were resolved through discussion among the authors.

Based on this mapping exercise, we asked two questions about each driver. First, we asked: *Do the SSP scenario narratives or extensions make explicit assumptions about this driver as part of the storyline itself?* We assigned drivers into six groups: drivers that are (1) explicitly described in the SSP narratives, or are (2) implicit in the SSP narratives based on reasonable inferences from other variables and broader trends; (3) drivers that have been explored qualitatively or quantitatively in SSP extensions, or are (4) implicit in those extensions; (5) drivers that are not represented in the SSPs as currently envisioned, but can be modeled under SSP scenarios; and (6) those that are not well aligned with the SSP framework.

Second, we asked: *Can researchers access or make quantitative future projections of this driver under the SSP scenarios?* We again assigned drivers into six categories: those that have already been quantified, either in (1) the core SSP quantifications, (2) as standard IAM variables, or (3) in other modeling work; (4) those that could be projected under the SSP scenarios, based on modeling with existing data and/or through scenario extension; (5) those that can be quantitatively modeled and projected into the future, but are misaligned with the SSP framework; and (6) those that are not readily quantified.

### Mapping of pathogen-specific drivers

To illustrate how different pathogen systems respond to distinct combinations of drivers across the pandemic emergence process, we developed qualitative vignettes for a small number of representative pathogens (Influenza A virus, MERS- and SARS-like coronaviruses, mpox virus, paramyxoviruses (e.g., Nipah virus), and simian retroviruses (e.g., HIV)) (**Table 1**). These pathogens were selected to span diverse transmission ecologies, spillover pathways, transmission modes (e.g., respiratory or sexually-transmitted), and public health contexts. For a subset of representative pandemic risk drivers identified in our purposive review, we qualitatively assessed the strength of evidence linking each driver to each pathogen system using four categories: strong evidence (++), moderate evidence (+), speculative or uncertain evidence (?), and not relevant (—). These classifications represent a narrative synthesis of the peer-reviewed disease ecology, epidemiology, and pandemic preparedness literature and are intended to illustrate broad differences among pathogen systems, not to provide a formal quantitative ranking. We additionally summarized the expected direction of change for each driver under the SSPs using four qualitative categories: increasing in most SSPs (↑), decreasing in most SSPs (↓), diverging substantially among SSPs (↕), or not well represented within the SSP framework (○). Together, these qualitative assessments illustrate how pathogen-specific driver profiles interact with projected socioeconomic futures and highlight where the current SSP framework aligns more or less with different components of pandemic risk.

**Table 1.**
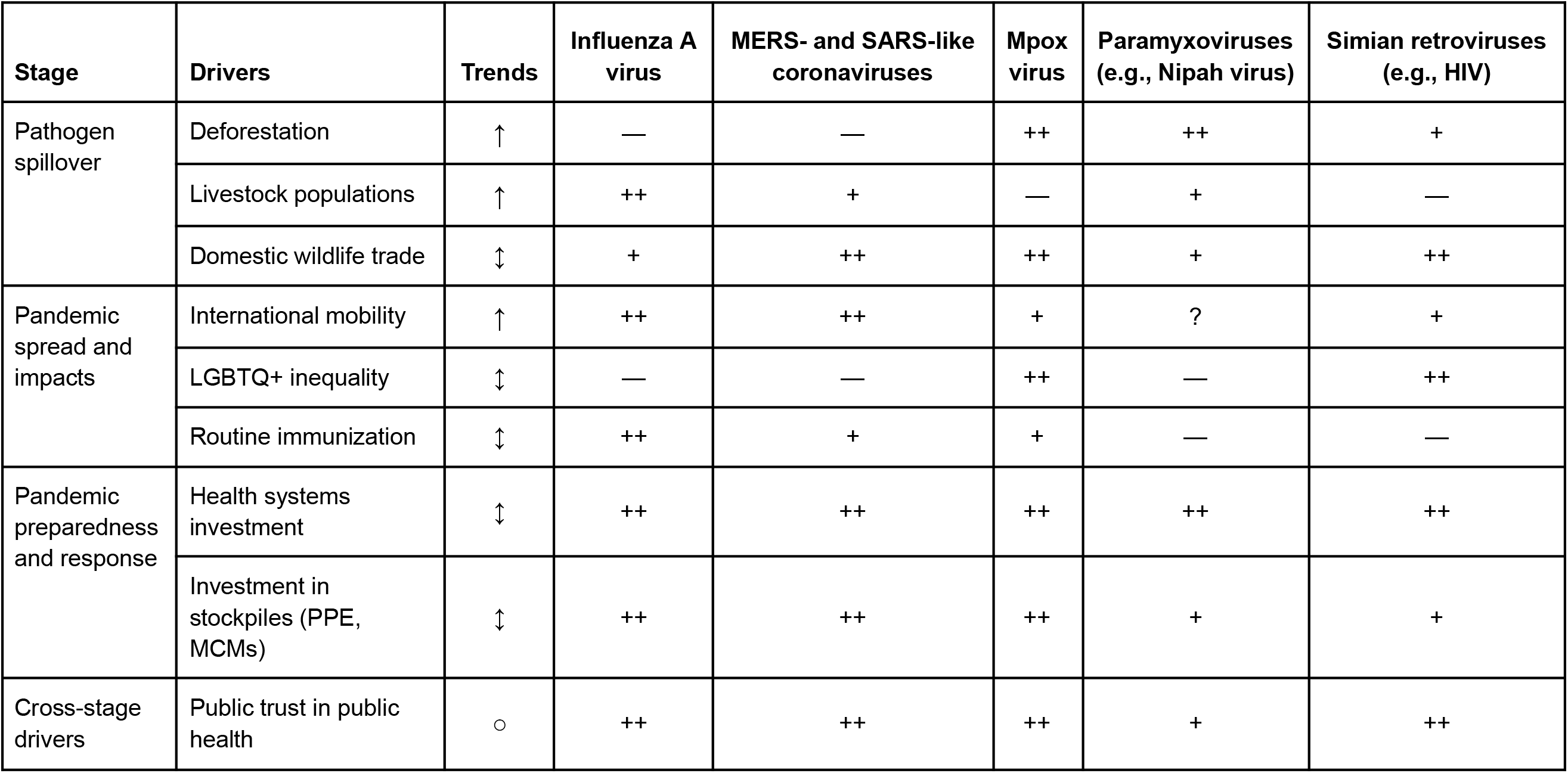
Representative pandemic pathogens and selected drivers across the pandemic emergence process, and projected trajectories of those drivers under the SSPs. Representative pathogen systems are compared according to the strength of evidence linking selected pandemic risk drivers to each disease system and the expected direction of change in those drivers across SSP futures. Driver–pathogen associations are shown as a qualitative synthesis of current evidence (strong (++), moderate (+), speculative (?), or not relevant (—)), while driver trajectories indicate whether each driver generally increases (↑), decreases (↓), diverges among scenarios (↕), or is not well represented within the SSP framework (○).

| Stage | Drivers | Trends | Influenza A virus | MERS- and SARS-like coronaviruses | Mpox virus | Paramyxoviruses (e.g., Nipah virus) | Simian retroviruses (e.g., HIV) |
| --- | --- | --- | --- | --- | --- | --- | --- |
| Pathogen spillover | Deforestation | ↑ | — | — | ++ | ++ | + |
|  | Livestock populations | ↑ | ++ | + | — | + | — |
|  | Domestic wildlife trade | ↕ | + | ++ | ++ | + | ++ |
| Pandemic spread and impacts | International mobility | ↑ | ++ | ++ | + | ? | + |
|  | LGBTQ+ inequality | ↕ | — | — | ++ | — | ++ |
|  | Routine immunization | ↕ | ++ | + | + | — | — |
| Pandemic preparedness and response | Health systems investment | ↕ | ++ | ++ | ++ | ++ | ++ |
|  | Investment in stockpiles (PPE, MCMs) | ↕ | ++ | ++ | ++ | + | + |
| Cross-stage drivers | Public trust in public health | ○ | ++ | ++ | ++ | + | ++ |

### Development of pandemic risk narratives

To synthesize our driver mapping results, we developed two complementary qualitative interpretations of the SSPs. First, we produced a set of pandemic-focused summaries of the five SSPs (**Box 2**). Second, we reinterpreted the original SSP scenario space from the perspective of pandemic risk by considering how each pathway is expected to influence opportunities for pathogen spillover and the subsequent spread and impacts of emerging pathogens (**Figure 4**). These products are not new scenario narratives or fully-developed extensions of the SSP framework, but instead recast the existing SSP storylines through the pandemic risk representation framework developed in this study.

To develop the new narratives, we began with the original SSP narrative paragraphs developed by O’Neill and colleagues^13^. We then synthesized the pandemic-relevant elements identified through our driver mapping exercise. Within each scenario summary, plain text describes the basic SSP storylines. Italicized text indicates our inferences based on the dominant themes of each storyline (i.e., implicit drivers), while bold text summarizes the overall implications for pandemic risk. For example, although SSP1 does not explicitly discuss pandemic vaccine distribution, its emphasis on multilateral cooperation, lower inequality, and stronger public institutions supports the inference that medical countermeasures would probably be developed and distributed more equitably during public health emergencies.

For the conceptual reinterpretation of the SSP scenario space, we summarized the narratives in **Box 2** and positioned each SSP in **Figure 4** according to two dimensions of pandemic risk: (i) the opportunities for pathogen spillover created by the ecological and socioeconomic conditions described by the scenario, and (ii) the likelihood that spillover events expand into devastating pandemics because of demographic, institutional, and public health factors. Whereas the original SSP scenario space is based on formal assumptions about barriers to climate change adaptation and mitigation, here we discuss consequences for pandemic risk as a more passive downstream impact of those same assumptions (versus as challenges to reducing risk).

### Code and data availability

Analyses were performed in the R statistical environment version 4.5.3. All data and code is available at https://github.com/carlsonlab/pandemic-ssp-public.

## Results

### The SSP framework captures spillover and spread better than response

First, we examined how comprehensively the SSP framework represents drivers acting across the pandemic emergence process (**Figure 1**, **Figure 2**). A clear pattern emerged: drivers acting early in the emergence process—particularly ecological conditions governing pathogen spillover risk—were substantially more likely to be explicitly represented in the SSP narratives and to have corresponding quantitative projections. In contrast, drivers associated with outbreak preparedness, response, and pandemic impacts were more frequently represented only implicitly, required SSP extensions or additional modeling, or remained difficult to quantify within the current framework.

**Figure 2.**
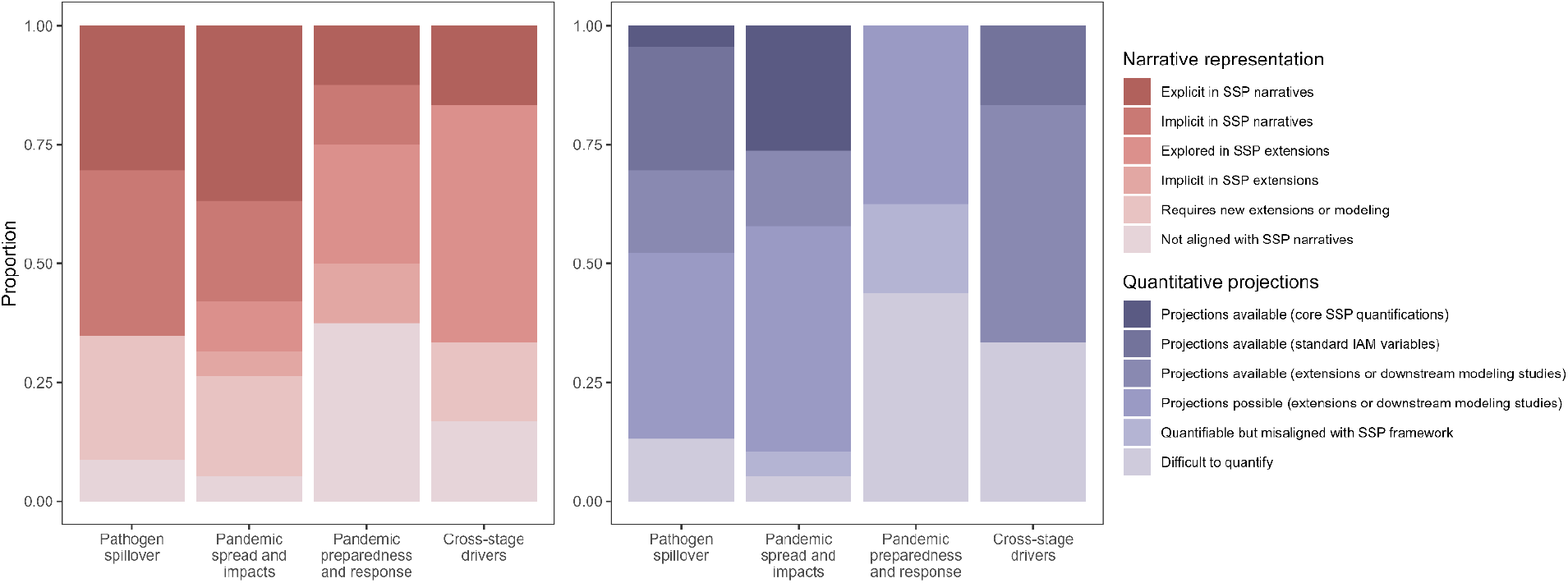
Pandemic risk driver representation within the SSP framework. Drivers are grouped by stages of pandemic risk (x-axis) and classified according to how they are represented within the SSP framework (see **Methods**; vertical groups).

The SSP scenarios are a reasonably good approximation of the socio-environmental trends driving changes in spillover risk. Drivers associated with land-use change, landscape fragmentation, livestock production systems, and dietary preferences are among the best represented components of the SSP framework, because they are central to analyses of land systems, agriculture, and greenhouse gas emissions, and have therefore been extensively quantified. However, the availability of these projections does not necessarily imply that they are equally well suited for downstream epidemiological applications; for example, translating coarse and highly uncertain land-use projections into ecologically realistic estimates of spillover risk remains an important challenge^45^. Furthermore, even within this stage, several important drivers—such as community awareness of wildlife-related risks, or laboratory biosafety and biosecurity—are not readily inferred from the existing narratives. These drivers are shaped less by long-term socioeconomic development than by evolving institutional, regulatory, and political decisions that can change rapidly over time and across governance scales.

Representation remains comparatively strong during the pandemic spread stage. Most variables widely used in models of human-to-human transmission—including population density, age structure, urbanization, mobility, and economic development—have been quantified within the SSP framework^27–29^. This includes a surprising degree of consideration of baseline population health: for example, the SSP narratives explicitly describe future investments in water, sanitation, hygiene, and health systems, and previous work has extended the framework to project non-communicable disease burdens under different socioeconomic futures^13,31^.

Coverage is weakest for the final stages of the pandemic emergence process: preparedness, response, and societal impacts. Although the SSP narratives (and health-related extensions^30,33^) describe broad differences in health systems, governance, surveillance capacity, migration, international cooperation, and institutional capacity, they provide only limited insight into the processes that often determine outbreak trajectories once a pathogen begins spreading. In particular, the framework provides relatively little information about public health decision-making, medical countermeasure innovation, emergency financing, or behavioral responses such as risk perception, vaccine acceptance, and adherence to public health recommendations. These processes are likely to depend not only on broader societal characteristics, including trust and social cohesion, but also on pathogen-specific factors such as disease severity and the perceived costs and benefits of intervention. Similarly, while the SSP narratives broadly characterize social inequality, they do not explicitly address several social and governance factors that shape equity in pandemic vulnerability, including barriers to medical countermeasure access and government propensity to restrict travel during outbreaks. Consequently, these downstream determinants of preparedness, response, and pandemic impacts are considerably less well articulated by the existing SSP framework than the upstream ecological and demographic drivers of spillover and transmission.

Taken together, these findings suggest that the SSP framework provides a useful foundation for exploring how global change shapes the ecological conditions of pathogen spillover and the epidemiological characteristics of pathogen transmission. Existing SSP projections also provide quantitative representations of many demographic, environmental, and socioeconomic drivers that could support simple scenario-based models of spillover and pandemic spread. However, these modeling efforts will be fundamentally limited, insofar as they need to address how societies detect, prepare for, and respond to outbreaks—and how this changes pandemic spread and impacts. Ongoing efforts to develop health-related extensions could close some of these gaps^33^, but these efforts are targeted at the health impacts of climate change instead of pandemic threats. As a result, drivers that are relevant to both of these health risks (e.g., health system capacity) will probably be better captured than those that are more specifically relevant to pandemic risk (e.g., public resistance to restrictive outbreak control measures).

These representation gaps will have different implications for different pathogen systems, which respond to distinct combinations of ecological, epidemiological, social, and institutional drivers across the pandemic emergence process (**Table 1**). For example, pandemic risks from influenza A viruses and MERS- and SARS-like coronaviruses are driven by livestock production and international mobility, both of which are comparatively well represented in the SSPs. In contrast, risks from mpox- and HIV-like pathogens are more shaped by dimensions of social vulnerability, such as inequality experienced by LGBTQ+ populations, that are less well-represented. These considerations will probably need to be made on a case-by-case basis for any given pathogen based on detailed knowledge of the relevant risk drivers at each stage of emergence.

### SSP scenarios imply diverging trajectories of pandemic risk

The SSPs describe substantially different futures for the ecological, demographic, economic, and institutional conditions that shape pandemic risk (**Figure 3 and Figure 4**; **Box 2**). Although the original narratives were developed to characterize socioeconomic challenges to climate change mitigation and adaptation, our synthesis suggests that they also imply distinct trajectories of risk across the entire pandemic emergence process. These trajectories arise because each SSP combines different assumptions about land use, agriculture, population dynamics, governance, inequality, and technological development, many of which correspond directly to the pandemic risk drivers identified in our mapping framework.

**Figure 3.**
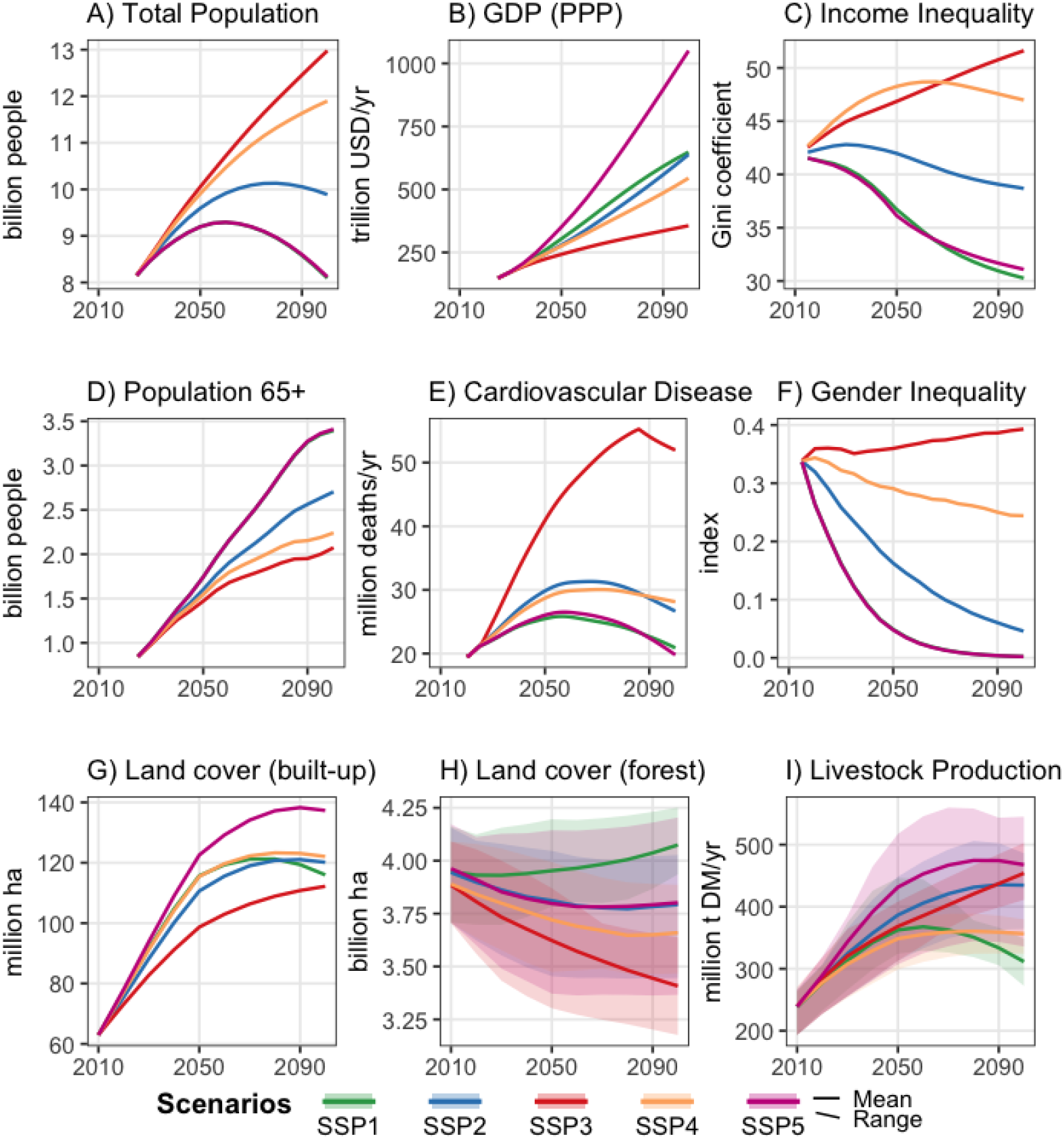
Projected trajectories of selected pandemic-relevant variables across the SSPs. Total population (A), age structure (D), and gross domestic product at purchasing-power parity (GDP-PPP) (B) are core quantifications of the SSP narratives. GDP values are reported in constant 2017 USD using the updated 2025 SSP basic-driver projections. Land cover (built-up) (G), land cover (forest) (H), and livestock production (I) are taken from IAM projections. In panel G, only one model (IMAGE) is shown; shaded ribbons in panels H and I indicate the range across IAM projections and are not statistical confidence intervals. Within-country income inequality (C)^54^, deaths from cardiovascular disease (E)^31^, and the Gender Inequality Index (F)^55^ are taken from SSP scenario extensions. Income inequality is a population weighted mean of national Gini index projections.

**Figure 4.**
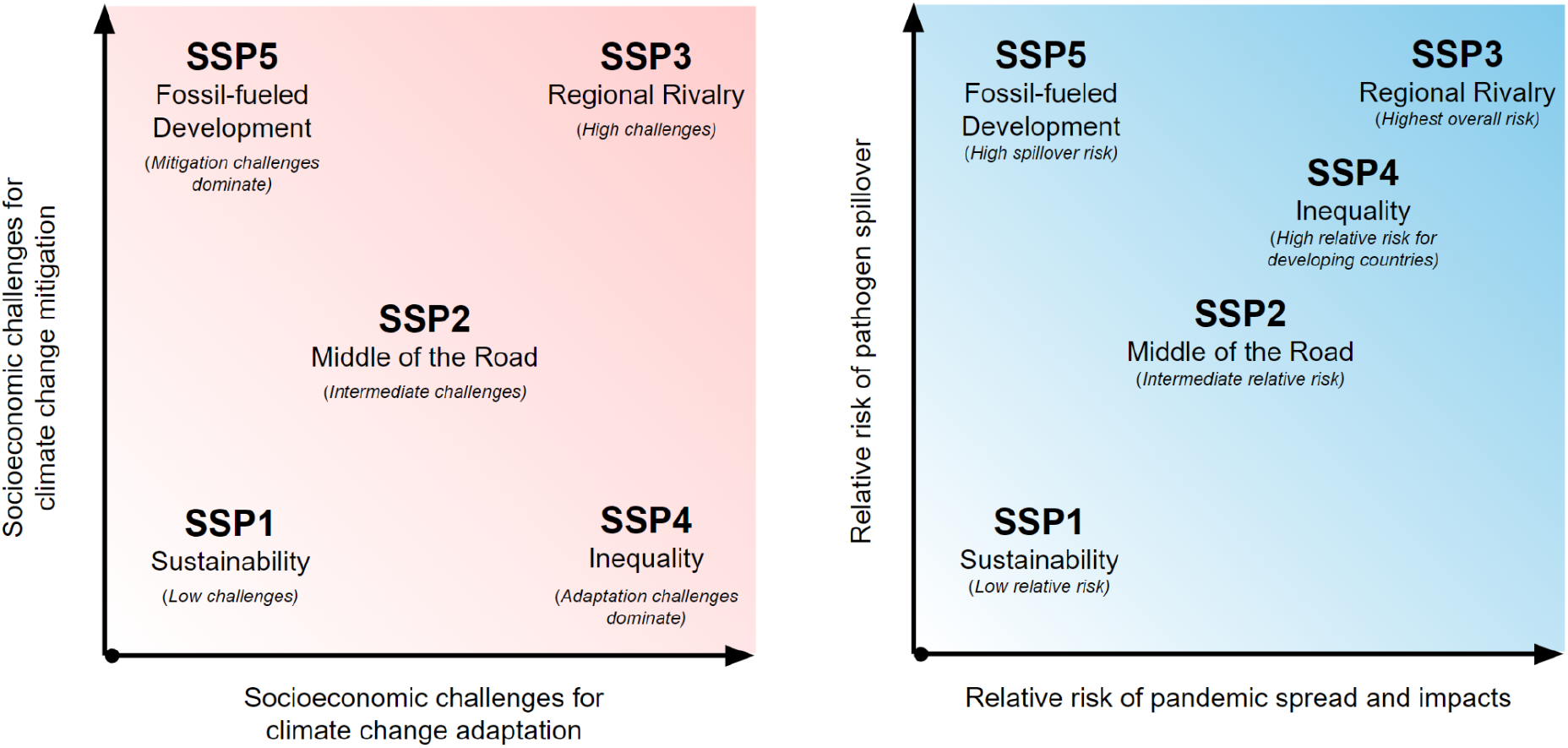
Translating the SSP scenario space into pandemic risk. Left: Original SSP scenario space adapted from O’Neill et al., in which the five SSPs are organized according to the challenges they pose for climate change mitigation (y-axis) and adaptation (x-axis)^12,13^. **Right:** Pandemic-risk interpretation developed in this study. Using the driver mapping presented in Figures 1–3, we reinterpret each SSP according to its implied opportunities for pathogen spillover (y-axis) and the likelihood that spillover events expand into pandemics with severe impacts (x-axis). Scenario placement represents a qualitative synthesis of the original SSP narratives and their quantitative implementations and should not be interpreted as a formal quantitative classification.

##### Box 2. “Pandemic risk narratives” for the SSPs.

Below, each paragraph’s plain text describes the basic SSP storylines. Italics indicate our inferences about pandemic drivers that are not explicit in the original narratives; bold indicates our summary of impacts on pandemic risk.

**SSP 1 (“Sustainability”):** The world prioritizes sustainable development, to the benefit of planetary health. Land and wildlife use are strongly regulated; diets rely less on animal protein, and particularly carbon- and land-intensive protein sources like beef; *and conservation efforts aim to preserve intact, biodiverse ecosystems*. **Collectively, this shift reduces contact between people, livestock, and wildlife, lowering spillover risk.** At the same time, substantial investment in pathogen surveillance and scientific innovation—together with progress towards universal health coverage and multilateral cooperation led by strong international organizations—**enables early outbreak detection, rapid response, and equitable distribution of vaccines and other medical countermeasures, and rapid deployment of new diagnostics and surveillance technologies. Most societies also become less vulnerable to the social and economic impacts of epidemics,** due to reductions in poverty and structural inequality.

**SSP 2 (“Middle of the Road”):** Societies follow historical patterns for population and economic growth, with incremental but uneven development. Some ecological drivers of spillover, such as deforestation and intensive livestock production, persist but are partially mitigated by gradual improvements in land management and conservation. Health systems and surveillance technologies strengthen unevenly, *with moderate investments in diagnostics, genomic surveillance, outbreak detection, and response, but continued disparities across regions*. Global governance mechanisms exist but progress slowly, hampered by bureaucracy and uneven compliance. Inequality narrows only slowly, leaving *many vulnerable populations at heightened risk.* **Pandemic risk continues to grow, but at a slower pace than in fractured or competitive futures.**

**SSP 3 (“Regional Rivalry”):** Global fragmentation drives national trends: politics is driven by nationalism and regional competition. Countries pursue food and energy security with weak regulation, leading to substantial land conversion from forests and other intact ecosystems into intensive and extensive agriculture and unplanned settlements. Geopolitical tensions reduce international trade in livestock *and wildlife,* driving demand for domestic production. These changes drive widespread biodiversity loss **and heightened zoonotic spillover risk**. International cooperation collapses, economic divides between and within countries grow, and many health systems remain weak and underfunded. Population growth is highest in low-income countries with the weakest institutions, amplifying vulnerabilities. Many governments pursue authoritarian policies, *undermining science-based public health during emergencies, reducing investment in surveillance innovation and international data sharing, and relying instead on border closures and other restrictive measures.* **Outbreaks spread rapidly within fragile systems, with little capacity for global coordination, equitable vaccine access, or timely containment.**

**SSP 4 (“Inequality”):** Global inequality widens, creating stark contrasts between highly connected, wealthy populations and poorer, marginalized ones. In high-income regions, strong ecological protections *and reduced contact with wildlife* **lead to lower spillover risk.** However, in the low-income, high-biodiversity tropical countries *that have historically faced the greatest burden of zoonotic spillover*, unregulated land use, extensive agriculture, and reliance on wild animal protein **lead to higher spillover risk**. Health investments are heavily skewed: elite regions invest heavily in advanced surveillance, genomic sequencing, diagnostics, and rapid medical countermeasure development, while poorer regions lack basic healthcare, sanitation, laboratory capacity, and the ability to manufacture or purchase medical countermeasures. **Outbreaks in vulnerable regions spread unchecked, eventually threatening the global community–but impacts differ dramatically between rich and poor countries**.

**SSP 5 (“Fossil-fueled Development”):** Rapid economic growth and resource-intensive lifestyles dominate. Diets remain meat-heavy and industrial livestock systems expand substantially; ecological protections are secondary to economic growth, leading to some ongoing deforestation, *and international wildlife trade intensifies*. Across the board, **unsustainable development raises risks of zoonotic spillover**. Countries and people become wealthier, and more people live in cities, leading to some baseline improvements in population health. Strong investments in science, biotechnology, digital surveillance, pathogen genomics, and health systems create *powerful capacities for early outbreak detection, rapid medical countermeasure development, and outbreak response, while globally connected markets support efficient manufacturing and purchasing during public health emergencies*. **Spillover events remain common, but most outbreaks are detected and contained before they develop into pandemics, particularly in wealthy and globally connected regions**.

The greatest divergence among scenarios occurs for upstream spillover drivers (**Figure 3**). Forest loss and broader patterns of land conversion range from being tightly regulated—and in some regions reversed—in SSP1 to continuing at moderate (SSP2, SSP5) or high (SSP3, SSP4) rates. Similarly, livestock production increases for at least several decades under all pathways, although the magnitude of growth differs substantially among scenarios. These trends are directly relevant to pathogen spillover because land-use change, agricultural intensification, and human–animal contact are among the best-established ecological drivers of zoonotic emergence. Likewise, although wildlife trade is not explicitly represented within the SSP scenarios, assumptions about conservation policy, dietary transitions, and environmental governance imply broadly parallel trajectories. These findings suggest that the most sustainable scenarios are likely to modestly reduce opportunities for zoonotic spillover, whereas the least sustainable scenarios substantially amplify many of its principal ecological drivers^46,47^.

The implications of the SSP narratives become more complex during the pandemic spread phase (post-emergence). Several demographic and socioeconomic drivers diverge substantially across scenarios, particularly population growth, aging, inequality, and health system strength. Global population and economic output increase under all pathways during the first half of the century, but the magnitude of these changes differs considerably. Population growth, poverty, and weak governance are most pronounced in SSP3, whereas rapid economic development, urbanization, and technological progress characterize SSP5. The narratives likewise describe contrasting trajectories for universal health coverage, access to water and sanitation, and broader investments in public health, with the greatest improvements occurring in SSP1.

Collectively, these assumptions describe a continuum between futures characterized by younger and rapidly growing populations, persistent poverty, weak institutions, and limited health system capacity—particularly SSP3 and, in many respects, SSP4—and futures with lower fertility, longer life expectancy, greater wealth, and stronger public institutions (SSP1, and to a lesser extent SSP2 and SSP5). These differences are likely to influence both the probability that outbreaks develop into widespread epidemics and the severity of their health, social, and economic consequences. However, the implications of some demographic trends remain pathogen dependent. Older populations may experience substantially higher mortality for respiratory pathogens such as influenza and COVID-19, while lower average contact rates and less densely connected social networks could reduce transmission opportunities for some directly transmitted infections^48^.

Preparedness and response capacity are hardest to compare between SSP scenarios, because they are minimally represented compared to ecological or demographic drivers. The SSP narratives primarily characterize future institutional capacity through broad assumptions about governance, education, inequality, international cooperation, and public investment, with comparatively little attention to surveillance systems or pandemic preparedness. Consequently, the scenarios probably imply substantial differences in outbreak detection, response capacity, and access to medical countermeasures, but these differences are often indirect and require interpretation from the broader socioeconomic context. For example, SSP1 consistently describes conditions associated with strong international cooperation, robust public institutions, and equitable access to health services, whereas SSP3 and SSP4 depict fragmented governance, weaker institutions, and persistent inequalities that would likely constrain coordinated outbreak response.

### Climate risk and pandemic risk are usually closely aligned

Although the SSPs were originally developed to span contrasting challenges to climate change mitigation and adaptation, we find that these same socioeconomic futures also define distinct trajectories of pandemic risk (**Figure 4**). Although individual pathogens are more or less responsive to different combinations of drivers (**Table 1**), the broad socioeconomic conditions represented by the SSPs tend to influence many of these drivers simultaneously. Consequently, the scenarios define coherent trajectories of overall pandemic risk despite important differences in the ecology, transmission, and emergence pathways of individual pathogen systems.

Importantly, we find that the same social, economic, and institutional conditions that make climate mitigation and adaptation more difficult also tend to increase pandemic risk; conversely, investing in environmental sustainability, stronger institutions, and lower inequality tends to reduce both spillover risk and vulnerability to outbreak impacts. This gradient is most obvious in the scenarios that face consistently low (SSP1), medium (SSP2), and high (SSP3) climate policy challenges, which we find are likely to be accompanied by closely corresponding levels of spillover risk and, consequently, overall pandemic risk. The two remaining scenarios tell subtly different stories. In both SSP4 and SSP5, zoonotic spillover risks are likely to be moderate to high; wealthy populations continue to eat meat-rich diets, and tropical land and conservation policy are weak, leading to continued escalation of human–animal contact in hotspots of zoonotic spillover. In SSP4, these increases in spillover risk lead to corresponding escalation of pandemic risk, but when a pandemic hits, wealthy countries experience substantially less devastating impacts. In contrast, SSP5 describes a more unfamiliar world, pairing environmental degradation and high levels of resource consumption with strong investments in science, technology, and health systems. In SSP5, outbreaks rarely become pandemics, but zoonotic spillovers remain common, with their burden concentrated in poorer regions—perhaps becoming, essentially, neglected tropical diseases. This pattern arguably already exists for some pathogens, such as Lassa virus, Rift Valley fever virus, and Crimean-Congo hemorrhagic fever virus, which are simultaneously treated as epidemic threats under the global health security agenda while disproportionately affecting low-resource settings and remaining chronically underfunded in biomedical R&D. The future implied by SSP5 evokes recent critiques that global health governance reforms over-prioritize progress on pandemic preparedness and response, which will always be inequitable in their distribution and outcomes, and under-prioritize strategies that reduce the health burden of spillover in frontline communities^49^.

Interestingly, neither SSP4 nor SSP5 describe the missing “fourth corner” world (**Figure 4**) where spillover is rare but, once pandemics start, they spread easily and have devastating impacts. This world resembles the recent past: a century ago, human and livestock populations were smaller, ecosystems were less degraded, populations were less connected, virology and clinical medicine were still in their infancy, and governments had nothing that looked like modern pandemic preparedness and response plans. Pandemics were rare, but—as in the case of the 1918 influenza pandemic or the sixth cholera pandemic—devastating in terms of mortality, long-term economic losses, and cultural impact. Given the range of plausible trends in the drivers of zoonotic spillover, it seems implausible that the world could return to this baseline.

## Discussion

Our analysis suggests that the SSP framework already contains much of the conceptual and quantitative machinery needed to investigate how long-term global change may influence pandemic risk. Although originally developed as a tool for climate change modeling and policy, the SSP framework captures many of the ecological, demographic, and institutional processes that shape opportunities for pathogen spillover and onward transmission. Across the five pathways, changes in deforestation, agricultural intensification, demography, mobility, inequality, and governance all have diverging impacts on pandemic risk. As a result, the SSPs already provide a coherent set of alternative futures within which pandemic risk can be explored.

Without (or before) constructing an entirely new scenario framework from scratch, pandemic researchers can build upon an internationally recognized framework that has already been widely adopted across climate, biodiversity, and sustainability science. The ongoing evolution of the SSP framework—including new health-specific extensions—also demonstrates that these scenarios are living frameworks that continue to expand into new domains^33,50^.

These findings also reinforce a growing consensus in disease ecology: escalating pandemic risk should not be considered separately from climate change and biodiversity loss, but instead, as another manifestation of the same underlying polycrisis of social-ecological systems.

Sustainable land management reduces opportunities for pathogen spillover, while reduced inequality, stronger public sector institutions, investments in global public goods and technology transfer, and international cooperation, strengthen societal capacity to detect and respond to outbreaks. Both sets of interventions also reduce climate risks, but they act on different stages of the pandemic emergence process, and SSP5 illustrates that a society can invest heavily in the second while neglecting the first. Conversely, scenarios characterized by continued ecosystem degradation, political fragmentation, weak governance, and widening inequality tend to increase both vulnerability to climate change and pandemic risk. Importantly, these relationships emerge because many policy domains simultaneously govern multiple global challenges. Decisions about industry, infrastructure, and inequality are all foundational to biodiversity conservation, climate action, and pandemic risk reduction. This strengthens the case for integrated planning approaches and reinforces recent calls from nexus frameworks and polycrisis research to evaluate climate, biodiversity, and health together instead of as independent policy domains^51,52^.

Despite these strengths, our results should not be interpreted as demonstrating that existing SSP products are immediately suitable for quantitative pandemic modeling. Although many pandemic-relevant variables are quantified, these projections have been developed to support a fundamentally different modeling task, and have important limitations. For example, land use projections can capture coarse changes in habitat availability, but do not capture ecologically realistic estimates of wildlife community composition, species interactions, or human-animal contact patterns. Considerable uncertainty therefore remains regarding how these variables should be translated into quantitative estimates of spillover risk. Similarly, many off-the-shelf quantifications of the SSP scenarios only make regional- or national-scale projections, while many determinants of outbreak detection and response—including surveillance capacity, health service delivery, water and sanitation infrastructure, and institutional effectiveness—operate at subnational or municipal scales. Consequently, additional spatial disaggregation will likely be required before SSP projections can support many epidemiological applications, and model intercomparisons have shown that such disaggregation approaches are highly uncertain^45^.

Therefore, we suggest that the SSPs provide a valuable starting point rather than a complete solution for future pandemic scenario analysis. Health-specific extensions currently under development could substantially improve representation of health systems, governance, surveillance, and other downstream determinants of outbreak trajectories^33^. Likewise, regional SSP extensions may better capture locally relevant institutions, environmental conditions, and development pathways than globally harmonized scenarios^32^. However, some limitations extend beyond missing variables. The SSPs intentionally describe a relatively constrained range of plausible socioeconomic futures developed primarily to support climate research. They do not explicitly explore more transformative possibilities—including fundamentally different economic systems, governance arrangements, or normative visions of desirable futures—that may substantially alter pandemic risk^53^. Nor do they systematically represent future innovation in pandemic preparedness, including advances in surveillance technologies, rapid vaccine development platforms, international financing mechanisms, or reimagined intellectual property regimes. These limitations suggest that pandemic preparedness may ultimately benefit from dedicated scenario frameworks analogous to those currently used in the planetary sciences.

Rather than replacing the SSPs, such frameworks could build upon their socioeconomic foundations while incorporating potential trajectories for public health institutions, technological innovation, behavioral adaptation, and equity considerations more explicitly.

Pandemic risk will persist under every plausible socioeconomic future. For at least a few decades, human populations will continue to grow, cities will expand, animal agriculture will continue, and opportunities for zoonotic spillover are unlikely to disappear entirely. At the same time, improvements in surveillance, health systems, medical countermeasure development, and outbreak response—and the degree to which these services are equitably distributed within and between countries—may substantially reduce the probability that spillover events become global catastrophes. Understanding the balance between these competing trends will require quantitative scenario analyses that integrate ecological, epidemiological, institutional, and behavioral processes over long time horizons. The SSP framework provides an important foundation for this work by already representing many of the upstream drivers of pandemic emergence. However, realizing the full potential of long-term pandemic risk assessment will require continued development of health-specific scenario extensions—and ultimately, perhaps, new scenario frameworks designed explicitly around the pandemic emergence process itself.

## Acknowledgements

TEL, CAS, DJB, RG, ZO, SJR, TP, SNS, and CJC were directly supported by the National Science Foundation (NSF DBI 2021909, 2213854, 2515340). RG was also supported by The Royal Society (University Research Fellowship URF\R1\251820). SJR was additionally supported by NSF grant DBI 2412115 and 2622265 as part of the US NSF Center for Analysis and Prediction of Pandemic Expansion (APPEX). LPB was supported by Future Ecosystems For Africa programme at the University of the Witwatersrand in partnership with Oppenheimer Generations Research and Conservation, the Swedish Research Council FORMAS grants African Futures (2020-00670) and SURPRISES (2025-01113) and the European Union’s Horizon Europe research and innovation programme under grant agreement No. 101213933 (INSPIRI). SP gratefully acknowledges funding from the Austrian Science Fund (FWF), project 346 REMASS, doi: 10.55776/EFP5. This project also benefitted from discussions at the Yale Pandemic Risk Scenarios Workshop (September 19-20, 2025), which was supported by a grant from PAX *sapiens* and the National Science Foundation (NSF DBI 2515340).

## References

1. Jones, K. E. et al. Global trends in emerging infectious diseases. Nature 451, 990–993 (2008).

2. Smith, K. F. et al. Global rise in human infectious disease outbreaks. J. R. Soc. Interface 11, 20140950 (2014).

3. Baker, R. E. et al. Infectious disease in an era of global change. Nat. Rev. Microbiol. 20, 193–205 (2022).

4. Meadows, A. J., Stephenson, N., Madhav, N. K. & Oppenheim, B. Historical trends demonstrate a pattern of increasingly frequent and severe spillover events of high-consequence zoonotic viruses. BMJ Glob Health 8, (2023).

5. Carlson, C. J. et al. Pathogens and planetary change. Nat. Rev. Biodivers. 1, 32–49 (2025).

6. Gibb, R., Franklinos, L. H. V., Redding, D. W. & Jones, K. E. Ecosystem perspectives are needed to manage zoonotic risks in a changing climate. BMJ 371, m3389 (2020).

7. Van Vuuren, D. P. et al. The scenario model intercomparison project for CMIP7 (ScenarioMIP-CMIP7). Geosci. Model Dev. 19, 2627–2656 (2026).

8. Pereira, L. et al. Advancing a toolkit of diverse futures approaches for global environmental assessments. *Ecosyst*. People 17, 191–204 (2021).

9. Cork, S. et al. Exploring alternative futures in the Anthropocene. Annu. Rev. Environ. Resour. 48, 25–54 (2023).

10. Moss, R. H. et al. The next generation of scenarios for climate change research and assessment. Nature 463, 747–756 (2010).

11. van Vuuren, D. P. et al. The representative concentration pathways: an overview. Clim. Change 109, 5–31 (2011).

12. O’Neill, B. C. et al. A new scenario framework for climate change research: the concept of shared socioeconomic pathways. Clim. Change 122, 387–400 (2014).

13. O’Neill, B. C. et al. The roads ahead: Narratives for shared socioeconomic pathways describing world futures in the 21st century. Glob. Environ. Change 42, 169–180 (2017).

14. Ebi, K. L. et al. Burning embers: synthesis of the health risks of climate change. Environ. Res. Lett. 16, 044042 (2021).

15. Rocklöv, J., Huber, V., Bowen, K. & Paul, R. Taking globally consistent health impact projections to the next level. *Lancet Planet*. Health 5, e487–e493 (2021).

16. Weber, E., Downward, G. S., Ebi, K. L., Lucas, P. L. & van Vuuren, D. The use of environmental scenarios to project future health effects: a scoping review. *Lancet Planet*. Health 7, e611–e621 (2023).

17. Berrang-Ford, L. et al. Systematic mapping of global research on climate and health: a machine learning review. *Lancet Planet*. Health 5, e514–e525 (2021).

18. Redding, D. W., Gibb, R. & Jones, K. E. Ecological impacts of climate change will transform public health priorities for zoonotic and vector-borne disease. bioRxiv (2024) doi:10.1101/2024.02.09.24302575.

19. Carlson, C. J. et al. Climate change increases cross-species viral transmission risk. Nature 607, 555–562 (2022).

20. Gibb, R., et al. The anthropogenic fingerprint on emerging infectious diseases. bioRxiv (2024) doi:10.1101/2024.05.22.24307684.

21. Trebski, A., Gourlay, L., Gibb, R., Imirzian, N. & Redding, D. W. Climate sensitivity is widely but unevenly spread across zoonotic diseases. Proc. Natl. Acad. Sci. U. S. A. 122, e2422851122 (2025).

22. Eby, P. et al. Pathogen spillover driven by rapid changes in bat ecology. Nature 613, 340–344 (2023).

23. Hsiang, S. M., Burke, M. & Miguel, E. Quantifying the influence of climate on human conflict. Science 341, 1235367 (2013).

24. Burke, M. B., Miguel, E., Satyanath, S., Dykema, J. A. & Lobell, D. B. Warming increases the risk of civil war in Africa. Proc. Natl. Acad. Sci. U. S. A. 106, 20670–20674 (2009).

25. Phillips, C. A. et al. Compound climate risks in the COVID-19 pandemic. Nat. Clim. Chang. 10, 586–588 (2020).

26. Redding, D. W. et al. Impacts of environmental and socio-economic factors on emergence and epidemic potential of Ebola in Africa. Nat. Commun. 10, 4531 (2019).

27. Kc, S. & Lutz, W. The human core of the shared socioeconomic pathways: Population scenarios by age, sex and level of education for all countries to 2100. Glob. Environ. Change 42, 181–192 (2017).

28. Dellink, R., Chateau, J., Lanzi, E. & Magné, B. Long-term economic growth projections in the Shared Socioeconomic Pathways. Glob. Environ. Change 42, 200–214 (2017).

29. Jiang, L. & O’Neill, B. C. Global urbanization projections for the Shared Socioeconomic Pathways. Glob. Environ. Change 42, 193–199 (2017).

30. Sellers, S. & Ebi, K. L. Climate change and health under the shared socioeconomic pathway framework. Int. J. Environ. Res. Public Health 15, E3 (2017).

31. Sellers, S. Cause of death variation under the shared socioeconomic pathways. Clim. Change 163, 559–577 (2020).

32. Dellar, M. et al. Creating the dutch One Health Shared Socio-economic Pathways (SSPs). Reg. Environ. Change 24, (2024).

33. Green, C., et al. Navigating global health futures under the Shared Socioeconomic Pathways. Lancet Planetary Health in press (2026).

34. García-Peña, G. E. et al. Land-use change and rodent-borne diseases: hazards on the shared socioeconomic pathways. Philos. Trans. R. Soc. Lond. B Biol. Sci. 376, 20200362 (2021).

35. Plowright, R. K. et al. Ecological countermeasures to prevent pathogen spillover and subsequent pandemics. Nat. Commun. 15, 2577 (2024).

36. Wolfe, N. D., Dunavan, C. P. & Diamond, J. Origins of major human infectious diseases. Nature 447, 279–283 (2007).

37. Lloyd-Smith, J. O. et al. Epidemic dynamics at the human-animal interface. Science 326, 1362–1367 (2009).

38. Xexakis, G., et al. Narrative and quantitative analysis of democratic principles in the Shared Socioeconomic Pathways. NPJ Clim. Action 5, (2026).

39. Bauer, N. et al. Shared Socio-economic pathways of the energy sector – quantifying the narratives. Glob. Environ. Change 42, 316–330 (2017).

40. Daigneault, A. et al. Developing detailed shared socioeconomic pathway (SSP) narratives for the global forest sector. *J*. For. Res. 34, 4–45 (2019).

41. Haddaway, N. R. & Watson, M. J. On the benefits of systematic reviews for wildlife parasitology. Int. J. Parasitol. Parasites Wildl. 5, 184–191 (2016).

42. Haddaway, N. R. et al. Eight problems with literature reviews and how to fix them. *Nat*. Ecol. Evol. 4, 1582–1589 (2020).

43. Plowright, R. K. et al. Pathways to zoonotic spillover. Nat. Rev. Microbiol. 15, 502–510 (2017).

44. Blacksell, S. D. et al. Laboratory-acquired infections and pathogen escapes worldwide between 2000 and 2021: a scoping review. Lancet Microbe 5, e194–e202 (2024).

45. Alexander, P. et al. Assessing uncertainties in land cover projections. Glob. Chang. Biol. 23, 767–781 (2017).

46. Jones, B. A. et al. Zoonosis emergence linked to agricultural intensification and environmental change. Proc. Natl. Acad. Sci. U. S. A. 110, 8399–8404 (2013).

47. Rohr, J. R. et al. Emerging human infectious diseases and the links to global food production. Nat. Sustain. 2, 445–456 (2019).

48. O’Driscoll, M. et al. Age-specific mortality and immunity patterns of SARS-CoV-2. Nature 590, 140–145 (2021).

49. Vora, N. M. et al. Interventions to reduce risk for pathogen spillover and early disease spread to prevent outbreaks, epidemics, and pandemics. Emerg. Infect. Dis. 29, 1–9 (2023).

50. O’Neill, B. C. et al. The Scenario Model Intercomparison Project (ScenarioMIP) for CMIP6. Geosci. Model Dev. 9, 3461–3482 (2016).

51. IPBES. Assessment Report on the Interlinkages Among Biodiversity, Water, Food and Health. (2024).

52. Pfenning-Butterworth, A. et al. Interconnecting global threats: climate change, biodiversity loss, and infectious diseases. Lancet Planet Health 8, e270–e283 (2024).

53. Pereira, L. M. et al. Solving science conundrums in the climate-nature-equity polycrisis with integrated transformative scenarios. One Earth 9, 101710 (2026).

54. Rao, N. D., Sauer, P., Gidden, M. & Riahi, K. Income inequality projections for the Shared Socioeconomic Pathways (SSPs). Futures 105, 27–39 (2019).

55. Andrijevic, M., Crespo Cuaresma, J., Lissner, T., Thomas, A. & Schleussner, C.-F. Overcoming gender inequality for climate resilient development. Nat. Commun. 11, 6261 (2020).

